# Depressive Symptoms and Physical Activity Mediate the Adverse Effect of Pain on Functional Dependence in Patients With Arthritis: Evidence From the Canadian Longitudinal Study on Aging

**DOI:** 10.1101/2025.02.10.25322043

**Authors:** Miriam Goubran, Zachary M. van Allen, Martin Bilodeau, Matthieu P. Boisgontier

## Abstract

**Objective:** Arthritis is a chronic condition affecting hundreds of millions of people worldwide, often leading to pain and functional limitations. This study aimed to investigate the direct and indirect effects of pain on functional dependence in individuals with arthritis. Depressive symptoms and physical activity were examined as potential mediators of this relationship.

**Methods:** A total of 6972 participants with arthritis (4930 with osteoarthritis and 694 with rheumatoid arthritis) were included from the Canadian Longitudinal Study on Aging. Linear and logistic regression models were used to assess the relationships between baseline usual presence of pain or discomfort, depressive symptoms, physical activity, and functional dependence at follow-up.

**Results:** Baseline pain was positively associated with depressive symptoms (b = 0.356 [95% CI: 0.310 to 0.402]) and negatively associated with physical activity (b = −0.083 [95% CI: −0.125 to −0.042]). Functional dependence at follow-up was significantly predicted by baseline pain (log OR = 0.607 [95% CI: 0.261 to 0.952]), depressive symptoms (log OR = 0.358 [95% CI: 0.184 to 0.533]), and physical activity (log OR = −0.598 [95% CI: −0.818 to −0.378]). Mediation analysis showed that 23.3% of the total effect of pain on functional dependence was accounted for by the indirect effect through depressive symptoms (16.2%), physical activity (6.3%), and their serial combination (0.8%).

**Conclusions:** The presence of pain at baseline was associated with higher odds of functional dependence in basic and instrumental activities of daily living after a mean follow-up period of 6.3 years, with depressive symptoms and lower physical activity acting as mediators.

**Impact:** Our findings highlight the need for arthritis care to extend beyond pain management by incorporating strategies that address depressive symptoms and promote physical activity to preserve functional independence.

## INTRODUCTION

The global prevalence of arthritis, particularly osteoarthritis, has risen significantly in recent decades, with cases increasing by 137% from 256 million in 1990 to 607 million in 2021.^1^ If trends continue, projections indicate that 1 billion people will have osteoarthritis by 2050.^2^ As a result, the burden of arthritis on society will continue to grow, since pain, a primary symptom, has been shown to predict functional decline.^3-6^

The current study builds on two theoretical frameworks explaining the relationship between pain and functional dependence. The fear-avoidance model (Vlaeyen and Linton, 2012) describes how individuals who catastrophize pain may develop pain-related fear, leading to avoidance behaviors, physical disuse, depression, and disability.^7^ Similarly, the avoidance model, specifically applied to osteoarthritis, describes how pain and psychological distress can initiate a cascade starting with the avoidance of activities and leading to muscle weakness and activity limitations.^8^ More recently, meta-analytic evidence has further supported the association between fear of movement and reduced physical activity across clinical populations, including rheumatologic conditions such as arthritis.^9^ These models and results emphasize the role of psychological and behavioral variables in the pathway from pain to functional dependence. Informed by these frameworks, our study tested depressive symptoms and physical activity as mediators of the effect of pain on functional dependence in individuals with arthritis, using a longitudinal dataset (Fig. 1). This empirical approach aimed to clarify the extent to which these mediators operate in parallel to influence functional dependence.

**Figure 1.**
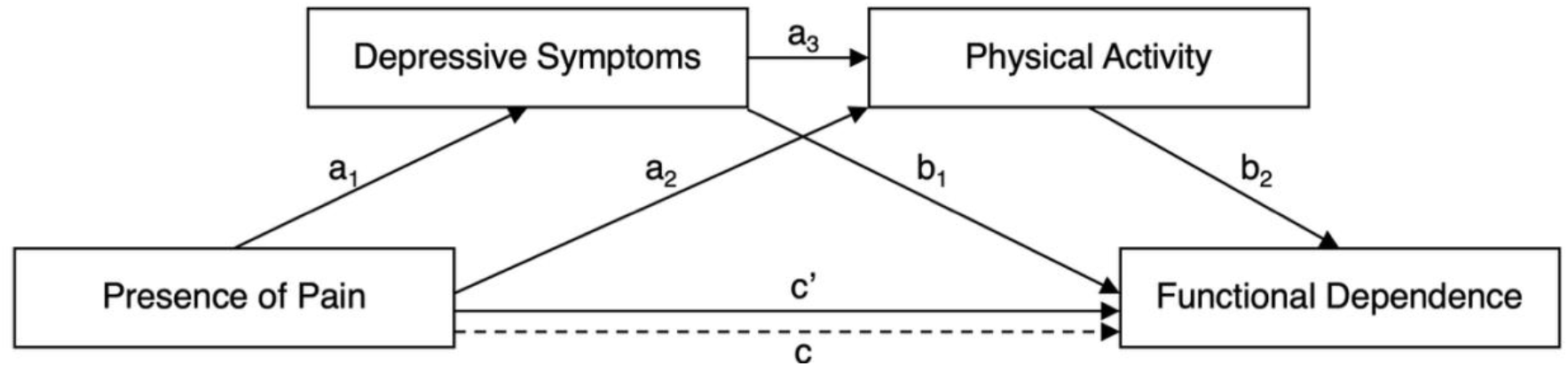
Path diagram of the serial mediation model testing the indirect effect of baseline pain on follow-up functional dependence via depressive symptoms and physical activity. c = total effect of pain when mediators are not adjusted for; c’ = direct effect of pain after adjusting for both mediators; a paths = effects of the independent variables on the mediators; b paths = effects of the mediators on the outcome variable, controlling for the independent variable; a_1_ × b_1_ = indirect effect through depressive symptoms; a_2_ × b_2_ = indirect effect through physical activity; a_1_ × a_3_ × b_2_ = indirect effect through depressive symptoms and physical activity. All paths are adjusted for age, sex, and baseline functional dependence.

Understanding the relationship between pain and functional dependence has important implications, as loss of independence significantly affects quality of life and healthcare costs.^10,11^ While pain directly contributes to functional decline,^3-6^ this effect may be mediated by factors related to mental and physical health. Two relevant candidate mediators of the relationship between pain and functional independence are depressive symptoms and physical activity, as both are influenced by pain^12-16^ and predict functional dependence.^6,17,18^ Understanding the complex relationships between pain, depressive symptoms, and physical activity, and how they collectively influence functional dependence in individuals with arthritis may inform targeted interventions in this population.

Depression is one of the most common comorbidities in arthritis, with recent meta-analyses suggesting that it affects approximately one-third of this population.^19,20^ The relationship between pain and depressive symptoms has been consistently reported in the literature^21^ and may be bidirectional.^22^ Longitudinal studies investigating osteoarthritis have shown that greater pain symptoms prospectively predict the incidence of depression,^23-25^ while other studies have shown that depressive symptomology may exacerbate the experience of pain.^26,27^ However, in the context of arthritis, the mechanistic explanations supporting the effect of pain on depression are stronger than those supporting the reverse, mainly because pain is a primary and more direct symptom resulting from joint inflammation, whereas depression is a comorbidity that develops secondary to the chronic pain.

The mediation of the effect of pain on functional dependence by depressive symptoms is well documented in the pain literature,^28-35^ particularly in studies on back pain^28-31^ and traumatic injury.^32-34^ However, to the best of our knowledge, this mediation has not been examined in people with arthritis. This is a notable gap, given that arthritis is one of the leading causes of chronic pain and disability. Understanding the mechanisms underlying the relationship between pain and functional dependence in this population may inform interventions aimed at improving functional outcomes.

In contrast to the proposed model (Fig. 1), studies in arthritis populations have primarily tested a different mediation model in which functional dependence is the mediator and depressive symptoms are the outcome,^36,37^ a model more relevant to mental health professionals. However, from a rehabilitation perspective, the focus is more on understanding the mechanisms that may improve functional outcomes. Moreover, conceptualizing depressive symptoms as a mediator aligns with the biopsychosocial approach to pain in rehabilitation,^38^ which emphasizes psychological contributions to physical functioning. The mechanistic basis for this mediation model is well supported, as depressive symptoms can reduce motivation,^39^ increase fatigue,^36,40,41^ and promote avoidance behaviors,^42^ all of which can affect physical function.^43^

Another potential mediator of the effect of pain on functional dependence is physical activity, as pain has been shown to predict physical activity levels,^44^ which in turn have been shown to improve physical functioning.^45^ While this indirect effect through physical activity has recently been examined in individuals with back pain,^46,47^ this mediating effect has not been examined in individuals with arthritis. As physical activity has been shown to be an effective intervention for improving pain and function,^48^ refining our understanding of this relationship, accounting for the influence of depressive symptoms, could further inform rehabilitation programs for patients with arthritis.

The aim of this study was to examine the relationship between pain and functional dependence in people with arthritis using the Canadian Longitudinal Study on Aging datasets. Specifically, we tested whether depressive symptoms and physical activity mediate the effect of baseline pain on functional dependence at follow-up (Fig. 1). We hypothesized that the presence of pain would have a direct effect on functional dependence. In addition, we hypothesized an indirect effect, whereby the presence of pain would be associated with higher depressive symptoms and lower physical activity levels, both of which would, in turn, predict higher odds of functional dependence, over and above the direct effect of pain.

## METHODS

### Participants

Participants in the Canadian Longitudinal Study on Aging (CLSA) were recruited by the CLSA team through Canada’s provincial health registries, random-digit dialing, and the Canadian Community Health Survey on Healthy Aging.^49,50^ Exclusion criteria for CLSA enrolment included residents living in Canada’s three territories and First Nations reserves, full-time members of the Canadian Armed Forces, individuals living with cognitive impairments, and individuals living in institutions, including 24-hour nursing homes.^49^ For the present study, we analyzed CLSA participants who had baseline measures of pain, depressive symptoms, physical activity, age, and sex, as well as measures of functional limitations at both baseline and follow-up, were included in the analyses. The study was approved by the University of Ottawa Research Ethics Board (H-10-23-9760).

Data of the ‘baseline’ assessment was collected between 2010 and 2015 using two approaches: data collection from a ‘tracking’ cohort of participants via 60-minute computer-assisted phone interviews and data collection from a ‘comprehensive’ cohort via 90-minute inperson interviews in addition to a data-collection site visit. Additionally, a ‘maintaining contact questionnaire’ was administered by phone to the comprehensive and tracking cohorts. The maintaining contact questionnaire, tracking cohort, and comprehensive cohort were used as baseline in our analyses. Between 2018 and 2021 another wave of data was collected during the follow-up assessment.

### Variables

#### Arthritis

At the baseline assessment, participants self-reported whether they had ever been diagnosed with rheumatoid arthritis, osteoarthritis, or any other type of arthritis. Five single-item questions were used: “Has a doctor ever told you that you have [osteoarthritis in the knee / osteoarthritis in the hip / osteoarthritis in one or both hands / rheumatoid arthritis / any other type of arthritis]?” Responses were yes or no. The dataset was initially filtered to include individuals with any type of arthritis for the main analyses. In addition, sensitivity analyses were conducted on two specific subsets: one comprising individuals with osteoarthritis and the other comprising those with rheumatoid arthritis.

#### Functional Dependence

Functional dependence in basic and instrumental activities of daily life (I/ADL) at baseline and follow-up was measured with a modified version of the Older Americans’ Resources and Services Multidimensional Functional Assessment Questionnaire (OARS).^51,52^ Twenty questions were asked to participants regarding their ability to complete seven basic activities of daily life (ADL) (e.g., can you eat without help, can you walk without help) in addition to 21 questions regarding seven instrumental activities of daily living (IADL) (e.g., can you use the telephone without help, can you prepare you own meals without help). The CLSA dataset includes a derived ordinal variable, the ‘Basic and Instrumental Activities of Daily Living Classification’ that produces the following scores: 1 (no functional limitations), 2 (mild functional limitations), 3 (moderate functional limitations), 4 (severe functional limitations), and 5 (total functional limitations). This variable is derived from OARS responses using a method that assigns extra weight to the ability to prepare meals and aligns with variables used in Statistics Canada’s Canadian Community Health Survey.^53^ For our analyses, we computed a binary variable from this CLSA-derived classification at both baseline and follow-up. These variables classify participants with no or mild limitations (scores 1 and 2) as functionally independent, while those with moderate, severe, or total limitations (scores 3 to 5) were classified as functionally dependent. This binary variable was used in the statistical models.

#### Pain

The usual presence of pain or discomfort was assessed with the question “Are you usually free of pain or discomfort?” Responses were yes or no. We acknowledge that this item may reflect intermittent or persistent pain rather than constant presence or absence of pain, and that it captures both pain and discomfort. However, for readability, we often refer to this variable as “pain”.

#### Depressive Symptoms

The Center for Epidemiological Studies Short Depression Scale (CESD-10)^54^ was used to assess depressive symptoms at baseline. The CESD-10 contains 10 items measuring depressive feelings, restless sleep, hopefulness for the future, and loneliness (e.g., “How often did you feel hopeful about the future?”; “How often did you feel that everything you did was an effort?”). Response options range from 3 [“All of the time (5-7days)”] to 0 [“Rarely or never (less than 1 day)”]. A sum score ranging from 0-30 was used in the analyses. When one response was missing, CLSA imputed the value using the mean of the remaining nine items to compute the sum score. Participants with more than one missing item were excluded from the analyses. Since the distribution for the CESD-10 was highly skewed, a log-transformed version of the variable was used in the analyses.

#### Physical Activity

Physical activity was measured at baseline with the Physical Activity Scale for the Elderly (PASE),^55^ which assesses the frequency of walking, light physical activity, moderate physical activity, strenuous physical activity, and exercise. Items asked participants to report on their activity levels over the previous 7 days on a 1 (never) to 4 (often, 5-7 days) scale and to indicate the time per day engaged in each of these activities on a 1 (less than 30 minutes) to 5 (four hours or more) scale. A total physical activity score was derived from these items according to the PASE administration and scoring instruction manual,^56^ resulting in scores from 0 to 485, with higher scores indicating higher levels of physical activity (see Suppl. Material 1 for scoring scheme).

### Statistical Analyses

All analyses were conducted in R version 4.4.1,^57^ and analysis scripts are publicly available.^58^ A serial mediation model, based on the process.R script (Model 6),^59^ was used to estimate total, direct, and indirect effects of baseline pain on functional dependence at follow-up. This regression-based approach^60^ was selected for its transparency and interpretability. To assess the robustness of the results, a sensitivity analysis based on structural equation modeling was conducted.

Functional dependence was assessed both at baseline (2011-2015) and at follow-up (2018-2021), while all the other variables were collected at baseline only. The independent variable was the usual presence of pain or discomfort (binary: 0 = usually free of pain, 1 = usually not free of pain), and the dependent variable was functional dependence at follow-up (binary: 0 = functionally independent, 1 = functionally dependent). The two mediating factors, physical activity and depressive symptoms, were measured as continuous variables. Untransformed values for the PASE (physical activity) and log-transformed values for CESD-10 (depressive symptoms) were standardized and included in statistical models in terms of standard deviations from their mean values in the cohort. All models were adjusted for age (standardized), sex (0 = female, 1 = male), and baseline functional dependence.

The total effect of pain on functional dependence (path c; Fig. 1) was estimated using a logistic regression model in which pain was the predictor, adjusted for control variables (age, sex, baseline functional dependence). The direct effect (path c’) was estimated using a logistic regression model that was adjusted for the control variables and also included both mediating variables. Coefficients for path a_1_, a_2_, and a_3_ were estimated as beta coefficients using linear regression. Coefficients for path b_1_ and b_2_ were estimated as log-odds coefficients (also known as logit coefficients or log odds ratios [log OR]) using logistic regression. The indirect effects were estimated using the product-of-coefficients approach. Because the outcome variable was binary, and in line with methodological recommendations,^59,61,62^ indirect effects were calculated on the log OR scale as the product of the beta coefficient from the linear regression (path a) and the untransformed coefficient from the logistic regression (path b). Interpretation of log ORs follows the convention for logistic regression: positive values indicate increased odds, and negative values indicate decreased odds. Because individual path coefficients are estimated from linear and logistic regression models that use linear and nonlinear link functions, the estimates of direct and indirect effects may not sum to equal the total effect on the log odds ratio scale. Statistical inference for the indirect effects was conducted via empirical bootstrapping with 5000 resamples. This approach is recommended for multivariate non-normal data as it provides adequate control of Type I error rates and confidence interval coverage.^63^ Confidence intervals (95% CI) that did not include zero were interpreted as evidence of statistical significance.

The indirect effect of pain on functional dependence was estimated as the sum of three indirect pathways (Figure 1): pain → depressive symptoms → functional dependence (a_1_ × b_1_); pain → physical activity → functional dependence (a_2_ × b_2_); and pain → depressive symptoms → physical activity → functional dependence (serial pathway; a_1_ × a_3_ × b_2_). Thus, the indirect effect of pain on functional dependence can be expressed as: Total indirect effect = (a_1_ × b_1_) + (a_2_ × b_2_) + (a_1_ × a_3_ × b_2_).

Statistical significance was assessed using a two-tailed α level of 0.05. The percentage of the total effect mediated by the mediators was calculated using the difference in log OR between the total effect (c) and the direct effect (c’), using the following formula: 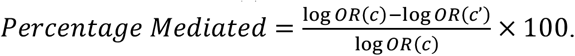 This calculation provides an alternative approach to quantifying mediation that does not rely on the individual components of the indirect effect described in the previous paragraph (i.e., the products of specific pathways shown in Figure 1).

### Sensitivity Analyses

Four sensitivity analyses were conducted to assess the robustness of our results. The first sensitivity analysis used a different binary classification of functional dependence at follow-up, classifying participants with no functional limitations as functionally independent and those with mild, moderate, severe, or total functional limitations as functionally dependent. In the second sensitivity analysis, the functional dependence variable was redefined to exclude meal preparation, which may be less relevant for participants with lower limb arthritis. The third and fourth analyses were stratified by arthritis type, focusing respectively on participants with osteoarthritis and those with rheumatoid arthritis.

### Confirmatory Analysis

To validate the main mediation model, we conducted a supplementary analysis applying structural equation modeling using the lavaan package,^64^ returning to the original variable definitions and categories. This confirmatory analysis replicated the main model’s pathways and estimated indirect effects using an alternative estimation method (Supplementary Material 2).

## RESULTS

### Descriptive Results

A total of 6972 participants reported having arthritis, including 4930 with osteoarthritis, and 694 with rheumatoid arthritis. Among the 6972 participants with any type of arthritis (Table 1), baseline data showed that 58.7% were female participants, more than half of them reported experiencing pain or discomfort (52.8%), the mean physical activity score was 185.0 ± 75.8, and the mean depression score was 5.5 ± 4.6. The majority (90.5%) reported no functional limitations, while 8.8% had mild limitations, and fewer than 1% had moderate or severe limitations. Most participants were aged 55-64 years (37.5%), followed by those aged 45-54 years (24.7%) and 65-74 years (24.4%), with the smallest proportion in the 75-85 age group (13.5%). The distribution of demographic and clinical characteristics for participants with osteoarthritis and rheumatoid arthritis is presented in Table 1. The average time between baseline and follow-up measures was 6.3 ± 0.8 years, with a range of 4.0 to 9.6 years.

**Table 1.** Sample characteristics at baseline by arthritis type.

|  | **Any Arthritis**<br>(n = 6972) | **Osteoarthritis**<br>(n = 4930) | **Rheumatoid Arthritis**<br>(n = 694) |
| --- | --- | --- | --- |
| **Variables** | Count (%) or<br>Mean (SD) | Count (%) or<br>Mean (SD) | Count (%) or<br>Mean (SD) |
| **Presence of Pain** |  |  |  |
| No Pain | 3288 (47.2%) | 2229 (45.2%) | 270 (38.9%) |
| Pain | 3684 (52.8%) | 2701 (54.8%) | 424 (61.1%) |
| **Physical Activity** | 185.0 (75.8) | 180.2 (76.0) | 190.9 (75.7) |
| **Depressive Symptoms** | 5.5 (4.6) | 5.5 (4.7) | 5.9 (4.8) |
| **Sex** |  |  |  |
| Male | 2882 (41.3%) | 1943 (39.4%) | 280 (40.3%) |
| Female | 4090 (58.7%) | 2987 (60.6%) | 414 (59.7%) |
| **Age Group** |  |  |  |
| 45-54 years | 1720 (24.7%) | 1002 (20.3%) | 207 (29.8%) |
| 55-64 years | 2612 (37.5%) | 1858 (37.7%) | 236 (34.0%) |
| 65-74 years | 1698 (24.4%) | 1332 (27.0%) | 169 (24.4%) |
| 75-85 years | 942 (13.5%) | 738 (15.0%) | 82 (11.8%) |
| **Functional Limitations** |  |  |  |
| None | 6310 (90.5%) | 4417 (89.6%) | 614 (88.5%) |
| Mild | 611 (8.8%) | 473 (9.6%) | 71 (10.2%) |
| Moderate | 43 (0.6%) | 34 (0.7%) | 5 (0.7%) |
| Severe | 5 (0.1%) | 4 (0.1%) | 2 (0.3%) |
| Total | 3 (0.05%) | 2 (0.05%) | 2 (0.3%) |

Notes: This table reports raw, unstandardized, and untransformed values for physical activity that were obtained from the PASE and values for depressive symptoms were obtained from the CESD-10. For use in regression models, raw PASE scores and log-transformed CESD-10 scores were standardized. The Functional Limitations variable reports counts of categories from the “Basic and Instrumental Activities of Daily Living Classification” derived in the CLSA dataset.

### Main Analyses

We used linear and logistic regression models to investigate the direct and indirect effects of baseline pain on functional dependence 6.3 ± 0.8 years later. Depressive symptoms and physical activity were examined as potential mediators. All analyses were adjusted for sex, age, and baseline functional dependence, with detailed results presented in Table 2. The main analyses were based on a sample 6972 participants with any type of arthritis.

**Table 2.** Regression coefficients and odds ratios for the mediation paths in the main serial mediation model (n = 6972)

| Variables | Path c<br>(outcome = functional dependence) |  |  | Path a<sub>1</sub><br>(outcome = depressive symptoms) |  |  |
| --- | --- | --- | --- | --- | --- | --- |
|  | Log OR | 95CI | p | b | 95CI | p |
| (Intercept) | -4.473 | -4.822; -4.150 | $<2 \times 10^{-16}$ | -0.137 | -0.176; -0.098 | $6.3 \times 10^{-12}$ |
| Age | 0.839 | 0.684; 0.997 | $<2 \times 10^{-16}$ | -0.072 | -0.095; -0.049 | $8.0 \times 10^{-10}$ |
| Sex | -0.291 | -0.625; 0.033 | 0.082 | -0.136 | -0.183; -0.089 | $1.1 \times 10^{-8}$ |
| Baseline Dependence | 3.439 | 2.810; 4.068 | $<2 \times 10^{-16}$ | 0.723 | 0.453; 0.992 | $1.5 \times 10^{-7}$ |
| Presence of Pain | **0.791** | **0.458; 1.138** | **$4.8 \times 10^{-6}$** | **0.356** | **0.310; 0.402** | **$<2 \times 10^{-16}$** |

| Variables | Path a<sub>2</sub> and a<sub>3</sub><br>(outcome = physical activity) |  |  | Path b<sub>1</sub>, b<sub>2</sub>, and c'<br>(outcome = functional dependence) |  |  |
| --- | --- | --- | --- | --- | --- | --- |
|  | b | 95CI | p | Log OR | 95CI | p |
| (Intercept) | -0.116 | -0.151; -0.082 | $5.2 \times 10^{-11}$ | -4.684 | -5.042; -4.326 | $<2 \times 10^{-16}$ |
| Age | -0.450 | -0.470; -0.430 | $<2 \times 10^{-16}$ | 0.640 | 0.465; 0.814 | $6.6 \times 10^{-13}$ |
| Sex | 0.401 | 0.360; 0.443 | $<2 \times 10^{-16}$ | -0.009 | -0.348; 0.329 | 0.958 |
| Baseline Dependence | -0.744 | -0.983; -0.505 | $1.1 \times 10^{-9}$ | 2.886 | 2.256; 3.516 | $<2 \times 10^{-16}$ |
| Presence of Pain | **-0.083** | **-0.125; -0.042** | **$8.0 \times 10^{-5}$** | **0.607** | **0.261; 0.952** | **$5.8 \times 10^{-4}$** |
| Depressive Symptoms | **-0.030** | **-0.050; -0.009** | **$5.3 \times 10^{-3}$** | **0.358** | **0.184; 0.533** | **$5.7 \times 10^{-5}$** |
| Physical Activity |  |  |  | **-0.598** | **-0.818; -0.378** | **$9.4 \times 10^{-8}$** |

Notes: b = regression coefficient (beta); log OR = log(Odds Ratio); 95CI = 95% confidence interval

#### Path c (Presence of Pain → Functional Dependence; without mediators)

Results showed a total effect (path c) of baseline pain on follow-up functional dependence (log OR = 0.791 [95% CI: 0.458 to 1.138]; P = 4.8 × 10^−6^) (Table 2, Fig. 2). This model explained 15.0% of the variance in functional dependence (McFadden’s R^2^ = 0.150) (Table 2, Fig. 2).

**Figure 2.**
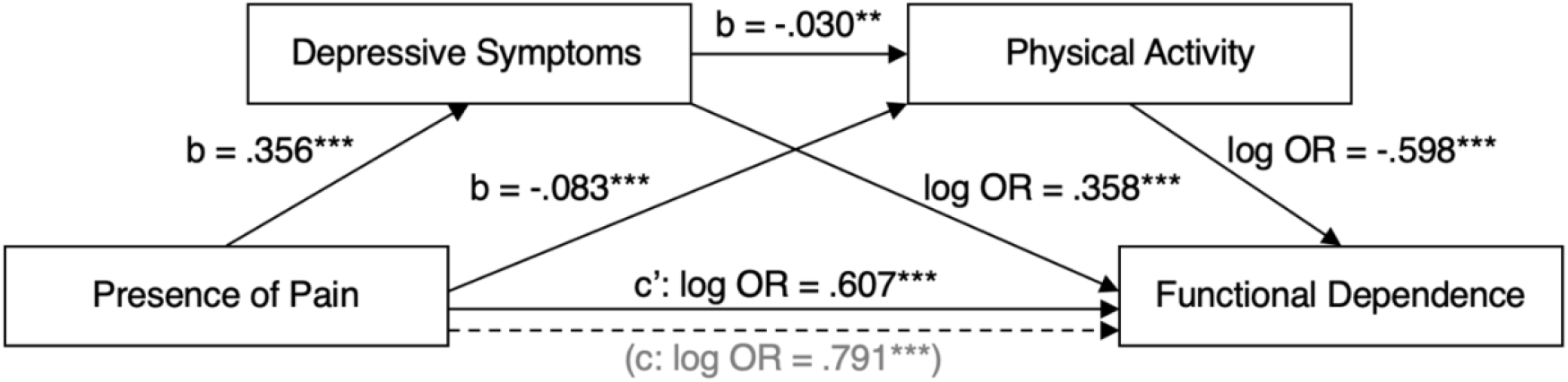
Path diagram displaying regression coefficients used to estimate direct and indirect effects of pain on functional dependence. b = beta, log OR = log(odds ratio), c = total effect, c’ (dash arrow) = direct effect, **p<.01, ***p<.001

#### Path a_1_ (Presence of Pain → Depressive Symptoms)

Results showed that within each category of age, sex, and baseline functional dependence, participants with pain usually present at baseline had log-transformed scores on the CESD-10 depression scale that were on average 0.356 standard deviations higher (95% CI: 0.310 to 0.402; *P* < 2.0 × 10^−16^) than participants usually free of pain (Table 2, Fig. 2). This model accounted for 4.7% of the variance in depressive symptoms (adjusted R^2^ = 0.047).

#### Path a_2_ (Presence of Pain → Physical Activity) and a3 (Depressive Symptoms → Physical Activity)

Participants with pain usually present at baseline engaged in less physical activity (path *a*_*2*_), with PASE scores that were on average 0.083 standard deviations lower (95% CI: –0.125 to – 0.042; *P* = 8.0 × 10^−5^) than participants usually free of pain. Similarly, participants with more depressive symptoms reported lower levels of physical activity (path a_3_). Specifically, participants who had log-transformed scores on the CESD-10 depression scale that were on average one standard deviation higher also had PASE physical activity scores that were on average 0.030 standard deviation lower (95% CI: –0.050 to –0.009; P = .005). Together, the variables in this model explained 25.3% of the variance in physical activity (adjusted R^2^ = 0.253) (Table 2, Fig. 2).

#### Path b1 (Depressive Symptoms → Functional Dependence), b2 (Physical Activity → Functional Dependence), and c’ (Presence of Pain → Functional Dependence, with both mediators)

Accounting for age, sex, and baseline functional dependence, higher depressive symptom scores were associated with higher odds of functional dependence. Specifically, each standard deviation increase in log-transformed scores on the CESD-10 depression scale was associated with a 0.358-unit increase (95% CI: 0.184 to 0.533; P = 5.7 × 10^−5^) in log odds of being functionally dependent at follow-up, 4.0 to 9.6 years later (Table 2, Fig. 2). Higher levels of physical activity at baseline were associated with lower odds of functional dependence at follow-up. Specifically, each standard deviation increase on the PASE was associated with a 0.598-unit decrease (95% CI: –0.818 to –0.378; P = 9.4 × 10^−8^) in the log odds of being functionally dependent (Table 2, Fig. 2).

Results showed a direct effect of baseline pain on functional dependence at follow-up (log OR = 0.607 [95% CI: 0.261 to 0.952]; *P* = 5.8 × 10^−4^) (Table 2, Fig. 2), indicating that participants with pain at baseline had higher log-odds of being functionally dependent 4.0 to 9.6 years later compared to those without pain. This model accounted for 18.0% of the variance in functional dependence (McFadden’s R^2^ = 0.180).

#### Indirect Effect

Results based on the product of the path coefficients (a × b) showed an indirect effect of baseline pain on functional dependence at follow-up (log OR = 0.184 [95% CI: 0.111 to 0.265]), which was partitioned into three indirect pathways through depressive symptoms (log OR = 0.128 [95% CI: 0.064 to 0.200]), physical activity (log OR = 0.050 [95% CI: 0.019 to 0.090]), and their serial combination (log OR = 0.006 [95% CI: 0.001 to 0.013]) (Fig. 3). The serial pathway with physical activity as the first mediator and depressive symptoms as the second mediator was also examined and yielded similar results (log OR = 0.0013 [95% CI: 0.0003 to 0.0028]). The exclusion of zero from all bootstrapped 95% CI indicated that each indirect effect was statistically significant.

**Figure 3.**
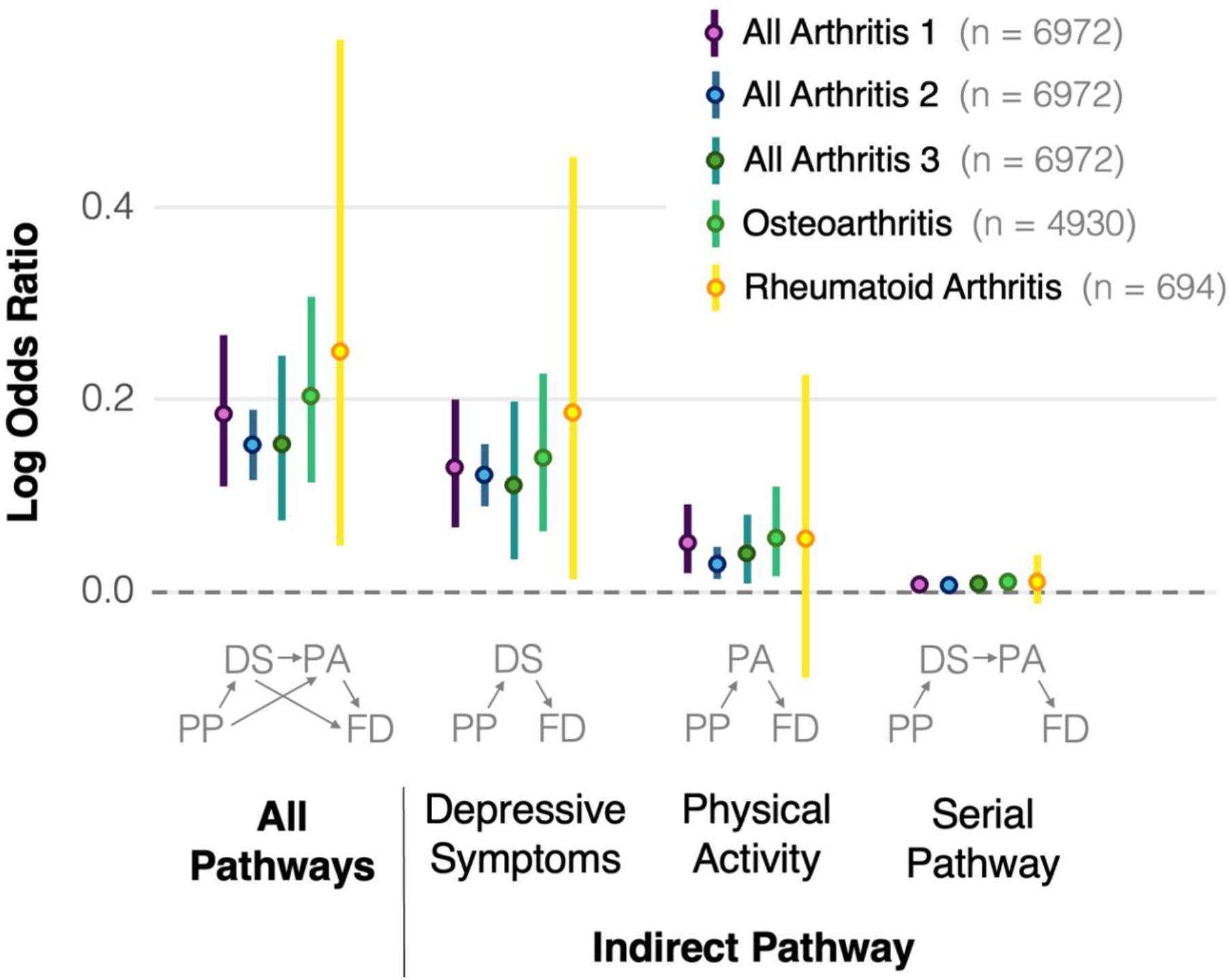
Indirect effect of baseline presence of pain on follow-up functional dependence. The indirect effect is partitioned into three indirect pathways through depressive symptoms (PP→DS→FD), physical activity (PP→PA→FD), and their serial combination (PP→DS→PA→FD). The horizontal dotted line at 0 indicates no effect, the dots represent estimated log odds ratios, and the vertical lines represent the 95% confidence intervals. Indirect pathways are presented for the main analysis, which included participants with any type of arthritis and classified them as functionally independent if they had no or mild functional limitations, and as functionally dependent if they had moderate, severe, or total functional limitations (All Arthritis 1). Four sensitivity analyses are also shown: participants with no functional limitations classified as functionally independent and those with mild, moderate, severe, or total functional limitations as functionally dependent (All Arthritis 2), measure functional dependence excluding meal preparation (All Arthritis 3), participants with osteoarthritis, and participants with rheumatoid arthritis. PP = presence of pain, DS = depressive symptoms, PA = physical activity, FD = functional dependence.

The comparison of the total effect (path c) and the direct effect (path c’) showed that 23.3% of the total effect of baseline pain on functional dependence was mediated by the indirect effect through the mediators: 16.2% through depressive symptoms, 6.3% through physical activity, and 0.8% through the serial mediation of depressive symptoms and physical activity.

### Sensitivity Analyses

#### Different Classification of Functional Dependence

The sensitivity analysis that classified participants with mild functional limitations as functionally independent rather than functionally dependent, yielded results similar to those of the main analysis. Results showed an indirect effect of baseline pain on functional dependence at follow-up (log OR= 0.152 [95% CI: 0.117 to 0.189]) (Fig. 3), which was partitioned into three indirect pathways: through depressive symptoms (log OR = 0.120 [95% CI: 0.089 to 0.154]), physical activity (log OR = 0.028 [95% CI: 0.013 to 0.046]), and their serial combination (log OR = 0.004 [95% CI: 0.001 to 0.007]).

#### Functional Dependence Excluding Meal Preparation

The sensitivity analysis based on a measure of functional dependence that excluded meal preparation yielded results similar to those of the main analysis. Results showed an indirect effect of baseline pain on functional dependence at follow-up (log OR= 0.154 [95% CI: 0.072 to 0.247]) (Fig. 3), which was partitioned into three indirect pathways: through depressive symptoms (log OR = 0.112 [95% CI: 0.036 to 0.199]), physical activity (log OR = 0.038 [95% CI: 0.009 to 0.079]), and their serial combination (log OR = 0.005 [95% CI: 0.001 to 0.011]) (Fig. 3).

#### Osteoarthritis

Results of the sensitivity analysis focusing on participants with osteoarthritis (n = 4930) were similar to those of the main analysis. Results showed an indirect effect of baseline pain on functional dependence at follow-up (log OR = 0.203 [95% CI: 0.113 to 0.307]), which was partitioned into three indirect pathways: through depressive symptoms (log OR = 0.139 [95% CI: 0.062 to 0.227]), physical activity (log OR = 0.055 [95% CI: 0.016 to 0.109]), and their serial combination (log OR = 0.009 [95% CI: 0.002 to 0.019]) (Fig. 3).

#### Rheumatoid Arthritis

Results of the sensitivity analysis focusing on participants with rheumatoid arthritis (n = 694) suggested that the total mediation (log OR = 0.250 [95% CI: 0.049 to 0.574]) was primarily driven by the pathway through depressive symptoms (log OR = 0.185 [95% CI: 0.013 to 0.453]) as the pathway through physical activity (log OR = 0.055 [95% CI: −0.089 to 0.226]) and the serial mediation pathway through both depressive symptoms and physical activity (log OR = 0.010 [95% CI: −0.013 to 0.038]) had bootstrapped 95% CI that included zero (Fig. 3), indicating that these indirect effects were not statistically significant.

### Confirmatory Analysis

A supplementary analysis using structural equation modeling (Suppl. Material 2) yielded results consistent with the main analysis in terms of the direction and significance of all paths. While estimates differed slightly in magnitude due to methodological differences, the overall pattern of direct and indirect effects was stable across approaches. These converging results further support the mediating roles of depressive symptoms and physical activity.

## DISCUSSION

### Main Findings

This study examined depressive symptoms and physical activity as potential mediators of the longitudinal relationship between baseline usual presence of pain or discomfort and functional dependence over a mean follow-up of 6.3 years in people with arthritis. Our results provide empirical evidence for a total effect of baseline pain on future functional dependence in ADLs and IADLs. Notably, 23.3% of this total effect was mediated by depressive symptoms and physical activity. However, as discussed earlier, estimates of direct and indirect effects may not sum exactly to the total effect, and this percentage should be interpreted as approximate. Consistent with the biopsychosocial approach to pain in rehabilitation,^38^ these findings highlight the importance of addressing both psychological and behavioral pathways in rehabilitation programs for patients with arthritis.

### Comparison With the Literature

Our results indicate that individuals experiencing pain at baseline had higher odds of functional dependence at follow-up, while adjusting for sex, age, and baseline functional dependence. This direct association between baseline pain and future functional dependence aligns with previous research demonstrating that pain is a key determinant of disability in individuals with arthritis.^3-6^ Taken together, these results suggests that early pain management may be critical in preventing long-term declines in functional abilities of individuals with arthritis.

Depressive symptoms were found to mediate the relationship between pain and functional dependence, accounting for 69.5% of the indirect effect. Specifically, our results suggest that individuals who reported being usually in pain or discomfort at baseline were more likely to experience depressive symptoms, which in turn increased the odds of functional dependence at follow-up. These results are consistent with those from studies of low back pain and post-traumatic pain that have documented the mediating role of depressive symptoms in the relationship between pain and functional dependence.^28-35^ Importantly, our findings extend this evidence to individuals with arthritis, highlighting the broader relevance of these mechanisms across pain-related conditions. As depression is both modifiable and treatable, these findings suggest that rehabilitation professionals should consider mental health when implementing interventions aimed at pain-related functional decline in individuals with arthritis.

Physical activity was also a mediator, accounting for 27.0% of the total indirect effect of pain on functional dependence. This finding is consistent with studies showing similar indirect effects in individuals with low back pain.^46,47^ Importantly, our findings extend this evidence to individuals with arthritis, reinforcing the importance of physical activity in maintaining functional independence in this population. The mediating effect is consistent with recent work in osteoarthritis showing that individuals with stronger tendencies to approach than avoid physical activity are less likely to reduce activity levels, even when experiencing high fear of movement.^65^ These results suggest that targeting both fear and motivational attitudes may improve physical activity and functional outcomes. Accordingly, exercise therapy should be considered as a component of rehabilitation programs aimed at restoring or preserving ADL and IADL independence in individuals with arthritis.

The mediation model we specified assumed a serial pathway in which depressive symptoms contribute to lower physical activity, which in turn affects functional dependence. The analysis yielded statistical evidence consistent with this hypothesized pathway. Although this serial effect accounted for only 3.5% of the total indirect effect, it suggests that individuals who develop depressive symptoms due to pain may subsequently reduce their physical activity, further exacerbating their risk of functional decline.

Our sensitivity analyses confirmed the robustness of our findings across different functional dependence classifications, statistical approaches, and arthritis subtypes. Particularly, analyses restricted to participants with osteoarthritis showed comparable mediation patterns, suggesting that the observed effects apply to the most common form of arthritis. However, in rheumatoid arthritis, the mediation effect was primarily driven by depressive symptoms, while the indirect effects through physical activity and the serial pathway were not statistically significant. This lack of significance may be due to the lower statistical power of this subsample but may also suggest that the mechanisms linking pain to functional decline differ between arthritis subtypes and warrant further investigation.

### Strengths and Limitations

This study has several strengths, including a large sample size of 6972 participants, a long follow-up period ranging from 4.0 to 9.6 years after baseline, the use of multiple mediation analyses, and bootstrapping to robustly estimate the significance of indirect effects. Several limitations should also be noted. First, the outcome and mediator variables were self-reported, which may have introduced measurement bias, especially in the context of a chronic condition like arthritis with a gradual onset. However, this limitation is common and often unavoidable in large cohort studies. Second, despite adjusting for key confounders, unmeasured variables such as medication use, disease severity, and comorbidities could have influenced the results. Third, since depressive symptoms and physical activity were both assessed at baseline, our mediation estimates may reflect concurrent associations rather than true causal pathways. Fourth, while CLSA has implemented sampling strategy designed to produce a nationally representative sample of adults aged 45 to 85 years at the time of recruitment, selection bias cannot be ruled out and may limit the generalizability of findings to individuals without access to a telephone, the internet, proficiency in English or French, or the ability to travel to a data collection site. Fifth, the operationalization of pain was based on a single binary item asking whether participants were usually free of pain or discomfort. This phrasing does not directly measure the presence, intensity, or duration of pain and may be subject to interpretation. Finally, although mediation analysis provides insight into potential causal pathways, the observational nature of the study precludes definitive causal conclusions.

### Conclusion

This study provides evidence suggesting that experiencing ongoing pain or discomfort contributes to long-term functional dependence both directly and indirectly through depressive symptoms and physical activity. The direct effect shows that individuals reporting pain have higher odds of being functionally dependent in ADLs and IADLs compared to those without pain. In addition, depressive symptoms and physical activity mediate this relationship.

Our findings underscore the need for a comprehensive approach to arthritis care that goes beyond pain management alone. They suggest that interventions should include strategies to address depressive symptoms and promote physical activity to mitigate the long-term impact of pain on functional dependence. For physical therapy practice, this means integrating routine screening for depressive symptoms into assessments and incorporating psychological support or referral pathways where needed. Furthermore, physical therapists should tailor exercise interventions to not only accommodate pain but also to enhance adherence and motivation, particularly in individuals exhibiting depressive symptoms, who may be at risk of physical inactivity. To support sustained engagement in physical activity, it is important to recognize that promoting health benefits alone is often insufficient.^66^ Instead, interventions should aim to foster pleasure and positive affective experiences related to physical activity.^67^

A biopsychosocial framework that integrates physical, psychological, and behavioral components may provide a more effective approach to maintaining, restoring, or improving physical function in individuals with arthritis. Clinically, this may involve interdisciplinary collaboration with mental health professionals and the use of behavior change techniques, such as goal setting, motivational interviewing, and graded activity, to promote physical activity despite pain.

## ARTICLE INFORMATION

### Author Contributions

Miriam Goubran: Conceptualization, Writing (Original Draft), Writing (Review and Editing); Zachary M. van Allen: Methodology, Data Curation, Formal Analysis, Writing (Original Draft), Writing (Review and Editing); Martin Bilodeau: Conceptualization, Writing (Review and Editing), Supervision (MG), Project Administration; Matthieu P. Boisgontier: Conceptualization (Lead), Methodology, Data Curation, Formal Analysis, Visualization, Writing (Original Draft) (Lead), Writing (Review and Editing), Supervision (MG and ZMvA), Project Administration, Funding Acquisition.

### Data and Code Sharing

In accordance with good research practices,^68^ the R scripts used to analyse the data are publicly available in Zenodo.^58^ This research was made possible using the data collected by the Canadian Longitudinal Study on Aging (CLSA) and was conducted using the CLSA Baseline Comprehensive Dataset version 7.0, Follow-Up 1 Comprehensive Dataset version 5.0, and Follow-Up 2 Comprehensive dataset version 2.0 under application number 2304015. This dataset is available for researchers who meet the criteria (www.clsa-elcv.ca). This manuscript was posted before peer review on the MedRxiv preprint repository on February 12, 2025.^69^

### Reporting Guidelines

This manuscript conforms to the AGReMA guidelines for mediation analyses of observational studies.^70^

### Disclaimer

The opinions expressed in this manuscript are the author’s own and do not reflect the views of the Canadian Longitudinal Study on Aging.

### Disclosure

The authors completed the ICMJE Form for Disclosure of Potential Conflicts of Interest and reported no conflicts of interest.

## SUPPLEMENTARY MATERIAL

**Supplementary Material 1. Scoring scheme for the Physical Activity Scale for the Elderly (PASE)**.

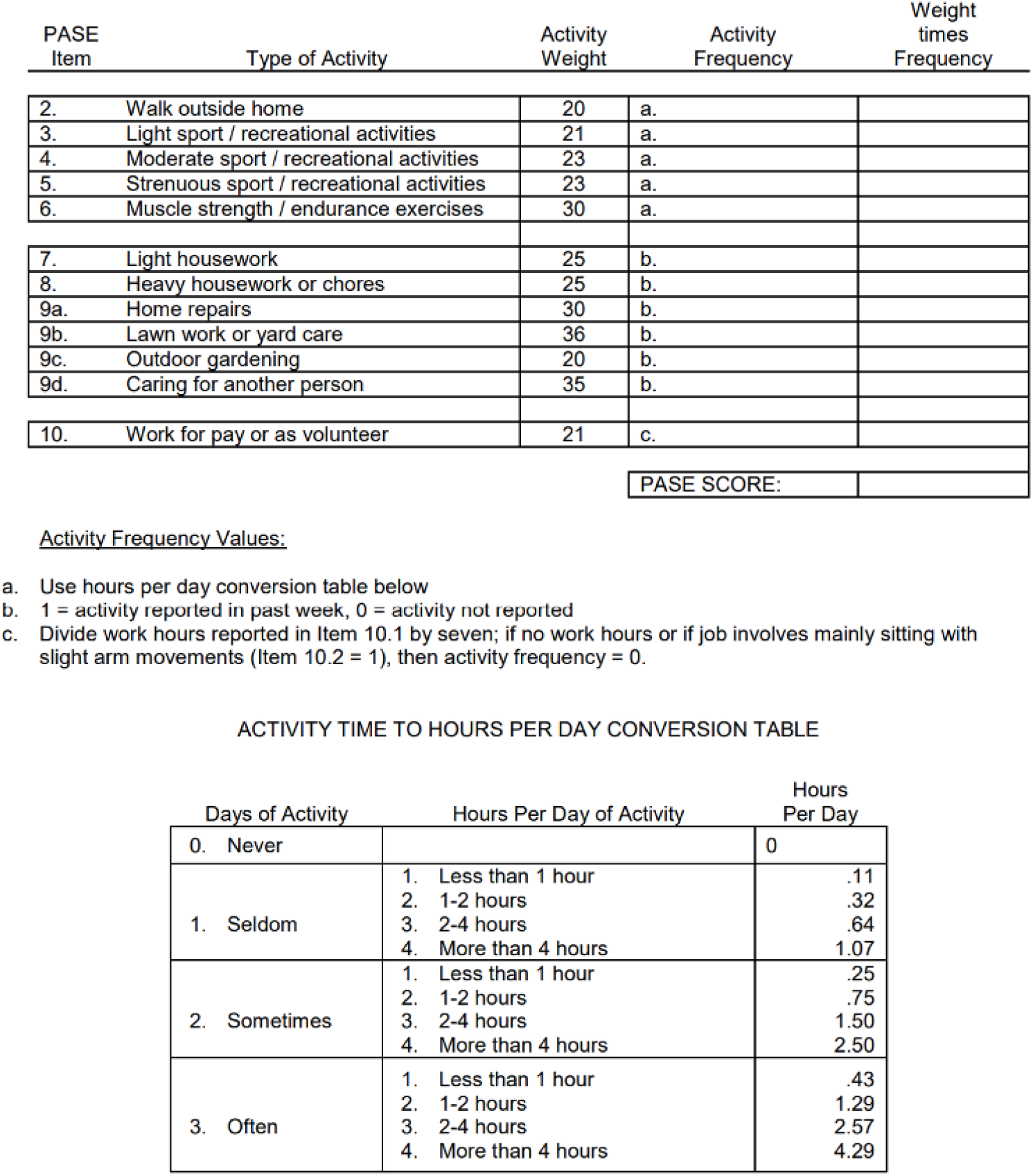

Full administration and scoring instruction manual available at https://meetinstrumentenzorg.nl/wp-content/uploads/instrumenten/PASE-handl.pdf

**Supplementary Material 2. Sensitivity analysis based on structural equation modeling (SEM)**

As a sensitivity analysis, we conducted a serial mediation model using structural equation modeling (SEM) with the *lavaan* package in R. The model specifies paths from pain to depressive symptoms and physical activity, with both mediators modeled as predictors of functional dependence at follow-up. Covariates included sex, age, and baseline functional dependence. The outcome was treated as an ordered categorical variable, and the model was estimated using the Weighted Least Squares Mean and Variance (WLSMV) estimator, which is appropriate for binary outcomes. Indirect effects were computed for three pathways: through depressive symptoms, through physical activity, and through the serial path.

Results showed that the direct effect of baseline pain on functional dependence at follow-up (c’) remained statistically significant after accounting for both mediators (log OR = 0.270, p < .001). In addition, the indirect effect of pain on functional dependence was statistically significant (log OR = 0.077, p < .001). This indirect effect was composed of three distinct pathways. The indirect effect through depressive symptoms alone was statistically significant (log OR = 0.056, p < .001), as was the indirect effect through physical activity alone (log OR = 0.018, p = .001). The serial pathway also yielded a small but significant indirect effect (log OR = 0.002, p = .008). The total effect of pain on functional dependence, combining both direct and indirect components, was estimated at log OR = 0.347 (p < .001). The model explained 4.7% of the variance in depressive symptoms, 25.4% of the variance in physical activity, and 21.3% of the variance in functional dependence.

**Suppl. Figure 1.**
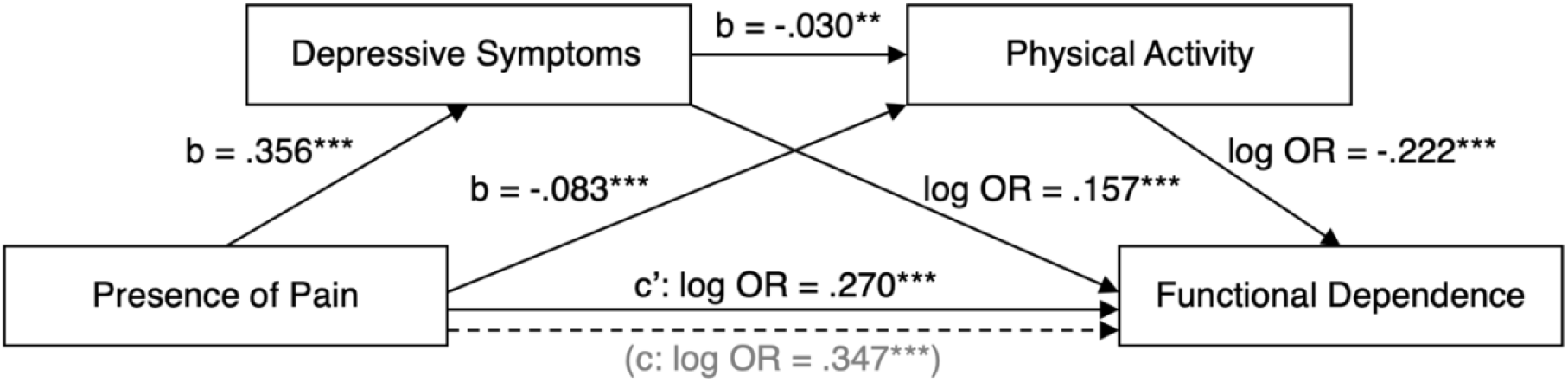
Serial multiple mediation model based on structural equation modeling using the *lavaan* package.

